# Disease waves of SARS-CoV-2 in Iran closely mirror global pandemic trends

**DOI:** 10.1101/2021.10.23.21265086

**Authors:** Zohreh Fattahi, Marzieh Mohseni, Maryam Beheshtian, Ali Jafarpour, Khadijeh Jalalvand, Fatemeh Keshavarzi, Hanieh Behravan, Fatemeh Ghodratpour, Farzane Zare Ashrafi, Marzieh Kalhor, Maryam Azad, Mahdieh Koshki, Azam Ghaziasadi, Mohamad Soveyzi, Alireza Abdollahi, Seyed Jalal Kiani, Angila Ataei-Pirkooh, Iman Rezaeiazhar, Farah Bokharaei-Salim, Mohammad Reza Haghshenas, Farhang Babamahmoodi, Zakiye Mokhames, Alireza Soleimani, Zohreh Elahi, Masood Ziaee, Davod Javanmard, Shokouh Ghafari, Akram Ezani, Alireza AnsariMoghaddam, Fariba Shahraki-Sanavi, Seyed Mohammad HashemiShahri, Azarakhsh Azaran, Farid Yousefi, Afagh Moattari, Mohsen Moghadami, Hamed Fakhim, Behrooz Ataei, Elahe Nasri, Vahdat Poortahmasebi, Mojtaba Varshochi, Ali Mojtahedi, Farid Jalilian, Mohammad khazeni, Abdolvahab Moradi, Alijan Tabarraei, Ahmad Piroozmand, Yousef Yahyapour, Masoumeh Bayani, Fatemeh Tavangar, Mahmood Yaghoubi, Fariba Keramat, Mahsa Tavakoli, Tahmineh Jalali, Mohammad Hassan Pouriayevali, Mostafa Salehi-Vaziri, Hamid Reza Khorram Khorshid, Reza Najafipour, Reza Malekzadeh, Kimia Kahrizi, Seyed Mohammad Jazayeri, Hossein Najmabadi

## Abstract

SARS-CoV-2 genome surveillance projects provide a good measure of transmission and monitor the circulating SARS-CoV-2 variants at regional and global scales. Iran is one of the most affected countries still involved with the virus circulating in at least five significant disease waves, as of September 2021. Complete genome sequencing of 50 viral isolates in an early phase of outbreak in Iran, shed light on the origins and circulating lineages at that time. As part of a genomic surveillance program, we provided an additional 319 complete genomes from October 2020 onwards. The current study is the report of complete SARS-CoV-2 genome sequences of Iran in the March 2020-May 2021 time interval. We aimed to characterize the genetic diversity of SARS-CoV-2 in Iran over one year.

Overall, 35 different lineages and 8 clades were detected. Temporal dynamics of the prominent SARS-CoV-2 clades/lineages circulating in Iran is comparable to the global perspective and introduces the 19A clade (B.4) dominating the first disease wave, followed by 20A (B.1.36), 20B (B.1.1.413), 20I (B.1.1.7) clades, dominating second, third and fourth disease waves, respectively. We observed a mixture of circulating 20A (B.1.36), 20B (B.1.1.413), 20I (B.1.1.7) clades in winter 2021, paralleled in a diminishing manner for 20A/20B and a growing rise for 20I, eventually prompting the 4^th^ outbreak peak. Furthermore, our study provides evidence on the entry of the Delta variant in April 2021, leading to the 5^th^ disease wave in summer 2021.

Three lineages are highlighted as hallmarks of SARS-CoV-2 outbreak in Iran; B4, dominating early periods of the epidemic, B.1.1.413 (specific B.1.1 lineage carrying a combination of [D138Y-S477N-D614G] spike mutations) in October 2020-February 2021, and the co-occurrence of [I100T-L699I] spike mutations in half of B.1.1.7 sequences mediating the fourth peak.

Continuous monthly monitoring of SARS-CoV-2 genome mutations led to the detection of 1577 distinct nucleotide mutations, in which the top recurrent mutations were D614G, P323L, R203K/G204R, 3037C>T, and 241C>T; the renowned combination of mutations in G and GH clades. The most frequent spike mutation is D614G followed by 13 other frequent mutations based on the prominent circulating lineages; B.1.1.7 (H69_V70del, Y144del, N501Y, A570D, P681H, T716I, S982A, D1118H, I100T, and L699I), B.1.1.413 (D138Y, S477N) and B.1.36 (I210del).

In brief, mutation surveillance in this study provided a real-time comprehensive picture of the SARS-CoV-2 mutation profile in Iran, which is beneficial for evaluating the magnitude of the epidemic and assessment of vaccine and therapeutic efficiency in this population.

## Introduction

Since December 2019, when the severe acute respiratory syndrome coronavirus 2 (SARS-CoV-2) was detected in Wuhan, China (Zhu et al. 2020), real-time whole-genome sequencing was begun to scrutinize the viral genome. Such genome surveillance projects aim to get the measure of transmission, manage the outbreak, monitor viral changes over time, and recognize the emerging viral variants (Oude Munnink et al. 2020). Alongside, broad and dynamic systems were designed to categorize the circulating SARS-CoV-2 variants raised by a large number of mutations; such as GISAID and Nextstrain nomenclature systems for major clades and a more dynamic algorithm for lineage classification called Pangolin (Shu and McCauley 2017, Hadfield et al. 2018, Rambaut et al. 2020). Based on the yearly mutation rate of 9.8×10^−4^ substitutions/site (Khateeb et al. 2021), multiple lineages/sub-lineages of SARS-CoV-2 have emerged from the two major A and B lineages at the root of SARS-CoV-2 phylogeny, and now 1293 Pango lineages, 12 Nextstrain and 9 GISAID clades are circulating worldwide (O’Toole et al. 2021). These lineages may be formed by one or a combination of deviant mutations (Khateeb et al. 2021). Among these, Variants of Concern (VOC) showed higher transmissibility and made severe waves of disease. As of September 2021, there have been four well-known VOCs; Alpha (20I clade, B.1.1.7 lineage), (Rambaut et al. 2020), Beta (20H clade, B.1.351 lineage) (Tegally et al. 2020), Gamma (20J clade, P.1 lineage) (Faria et al. 2021), and Delta (21A clade, B.1.617.2 lineage) (Mlcochova et al. 2021).

Thus far, different studies have monitored circulating SARS-CoV2 lineages at global and regional scales. In December 2020, Cella et al. studied 261,487 public genome sequences and reported B and B.1 as the most prevalent lineages in the world (Cella et al. 2021). Besides, 20I (B.1.1.7 lineage) was the major clade from February 2021 until 6 June based on the Nextstrain reports, when the 21A (B.1.617.2 lineage) gradually became dominant in the world. Such studies are also routinely performed at regional scales. For instance, analysis of 353 SARS-CoV-2 genomes in South-Eastern Italy introduced 20E, 20B, and 20A as the circulating clades in over one year with the dominancy of 20E (Capozzi et al. 2021). During November 2020-March 2021, another study in the United States sequenced 1600 SARS-CoV2 genomes. This study reported 20G, 20I, 20C, 20A, and 20B as the most predominant circulating clades and B.1.2, B.1.1.7, B.1.243, B.1.596, and B.1.526.1 as the most common lineages in National Capital Region (Morris et al. 2021).

Iran is one of the most affected Asian countries that confronted the spread of SARS-CoV-2 in at least five significant waves of SARS-CoV-2 infection, as of September 2021. The first COVID-19 cases were officially announced in February 2020, which soon led to the first outbreak peak from March continuing until Mid-May 2020. Then, whole-genome sequencing of 50 viral isolates from the early phase of the epidemic revealed B4 (19A clade) as the main SARS-CoV-2 lineage, while detected a surge of B.1.* lineages from May 2020 onward (Fattahi et al. 2021). Indeed, the country was confronted with the second increase in coronavirus patients/deaths from early June to the end of August 2020. The third disease wave gradually started in early September to the end of December 2020, with the highest number of positive COVID-19 tests reported on 28 November. Fortunately, a relative decrease in the number of patients was declared for the entire country from January 2021 to the end of March. However, insufficient restrictions during the holiday in March caused the emergence of a notable rise in the number of patients that led to the fourth outbreak peak started from April 2021 until early June. At that time, the highest rate of deaths since the beginning of the pandemic was recorded on 27 April 2021 (496 daily deaths). Less than two weeks later, the 5^th^ outbreak peak started from southern parts of Iran, expanded to the northern regions reaching 709 daily deaths on 25 August 2021. Overall, 5,295,786 cases and 114,311 deaths due to this disease are reported officially, as of 13 September 2021 (https://covid19.who.int/region/emro/country/ir). The current study is the report of complete genome sequences of SARS-CoV-2 viral isolates from Iran in the time interval of March 2020-May 2021. We aimed to characterize SARS-CoV-2 genetic diversity and circulating lineages/clades, especially those corresponding to the four infection waves of the disease. To accomplish this, we started with genome sequencing of 50 SARS-CoV-2 samples from the early phase of the epidemic (Fattahi et al. 2021) and then continued sequencing of additional 319 viral isolates. In summary, we provided a comprehensive picture of the SARS-CoV-2 epidemic in Iran; over one year after emerging.

## Methods

### Specimen recruitment, Sequencing and Genome assembly

As part of a genome surveillance program, 319 viral RNAs extracted from respiratory tract samples of laboratory-confirmed patients were collected from October 2020 till the end of May 2021 (TableS1). Most of the samples were collected randomly from different cities in Iran through the help of the Iranian Network for Research in Viral Diseases (INRVD) or from private laboratories. Moreover, a minor fraction of viral RNAs were referred from contact tracing and screening programs conducted in the Pasteur Institute of Iran (IPI). These samples were suspected of a special VOC. Whole-genome sequencing was performed following the CleanPlex® SARS-CoV-2 Research and Surveillance Panel (Paragon Genomics, Inc.) protocol. The targeted libraries were then paired-end sequenced on Illumina MiSeq instrument using 300-cycle MiSeq v2 reagent kits (Illumina, Inc.). On average, 98.8% of the SARS CoV-2 reference genome (NC_045512.2) was covered with an average depth of coverage of 1886.55X.

Initially, FASTQ files were assessed by FastQC (Andrews 2010) and then processed and compared with two in-house pipelines simultaneously. Fastp (Chen et al. 2018) and Cutadapt (Martin 2011) programs were applied for FASTQ pre-processing, followed by alignment to the NC_045512.2 using Bowtie2 (Langmead and Salzberg 2012) or Burrows-Wheeler Aligners (Li and Durbin 2009), keeping the high-quality reads mapped in proper pair. The filtered BAM files were used for assembly of consensus SARS-CoV-2 sequences with Samtools mpileup and Bcftools (Li et al. 2009). Finally, the consensus FASTQ files were converted into FASTA format by Seqtk (https://github.com/lh3/seqtk), masking bases with quality lower than 20 to ambiguous nucleotides (N). Furthermore, the samples with lower quality were re-checked in parallel using the automated Genome Detective system, trying to obtain the most accurate final FASTA file (Vilsker et al. 2019).

### Lineage and Clade assignment, Mutation analysis

To generate a comprehensive picture of SARS-CoV-2 genetic diversity in Iran, 50 FASTA files generated from the initial phase of this study (Fattahi et al. 2021), were added to this cohort and a total of 369 sequences remained for lineage and clade assignments using Pangolin v3.1.7 (Rambaut et al. 2020) and NextClade v.1.5.2 (Hadfield et al. 2018), respectively. As the samples were recruited in two separate projects, sampling in July-September 2020 interval was incomplete. Virus tracking in these months is accomplished using additional 14 samples submitted by other groups in GISAID (TableS2). Next, 25 sequences containing >5% ambiguous nucleotides (N) and bad overall NextClade QC score were removed from the cohort of samples subjected for mutation analysis. Detecting substitutions, deletions, and insertions were performed relative to NC_045512.2, using NextClade and Coronavirus typing tool (Cleemput et al. 2020).

## Results

### Cohort Description

In total, 369 SARS-CoV-2 samples were ascertained from patients with a history of COVID-19 symptoms and positive real-time RT-PCR assay. These samples were obtained from 19 different provinces during a time interval starting from 2 March 2020 until 27 May 2021 (Fig.1). The patient’s age ranged from 11 to 93 years old (Mean: 45.35 yrs.); 201 (54.5%) men (Mean: 44.38 yrs.), 168 women (Mean: 46.50 yrs.). Based on the severity of the disease, 136 (36.9%) recruited cases needed hospitalization, but 207 (56.1%) individuals were outpatients at the time of sample collecting (TableS1.). This cohort comprises at least 10 samples monthly and covers nearly the four peaks of SARS-CoV-2 spread in Iran; 188 (51%) of the samples were obtained during the outbreak peaks, and the other half were collected to track the viral changes in time intervals flanked by the outbreak peaks. The following time intervals were considered for the four disease waves as follow; First peak (19 February 2020-15 May 2020), Second peak; summer 2020 (1 June 2020 - 31 August 2020), Third peak; autumn 2020 (1 September 2020 – 1 January 2021), Fourth peak; spring 2021 (1 April 2021-June 2021) based on WHO Coronavirus (COVID-19) Dashboard (https://covid19.who.int/region/emro/country/ir). The number and percent of samples taken from each city, each month and each outbreak peak are shown in Fig.1A-B. The observed inflation in the number of samples ascertained in January 2021 (103 (27.9%)) is due to the need for tracking the possible entry of the Alpha variant into the country at that critical time.

**Figure 1.**
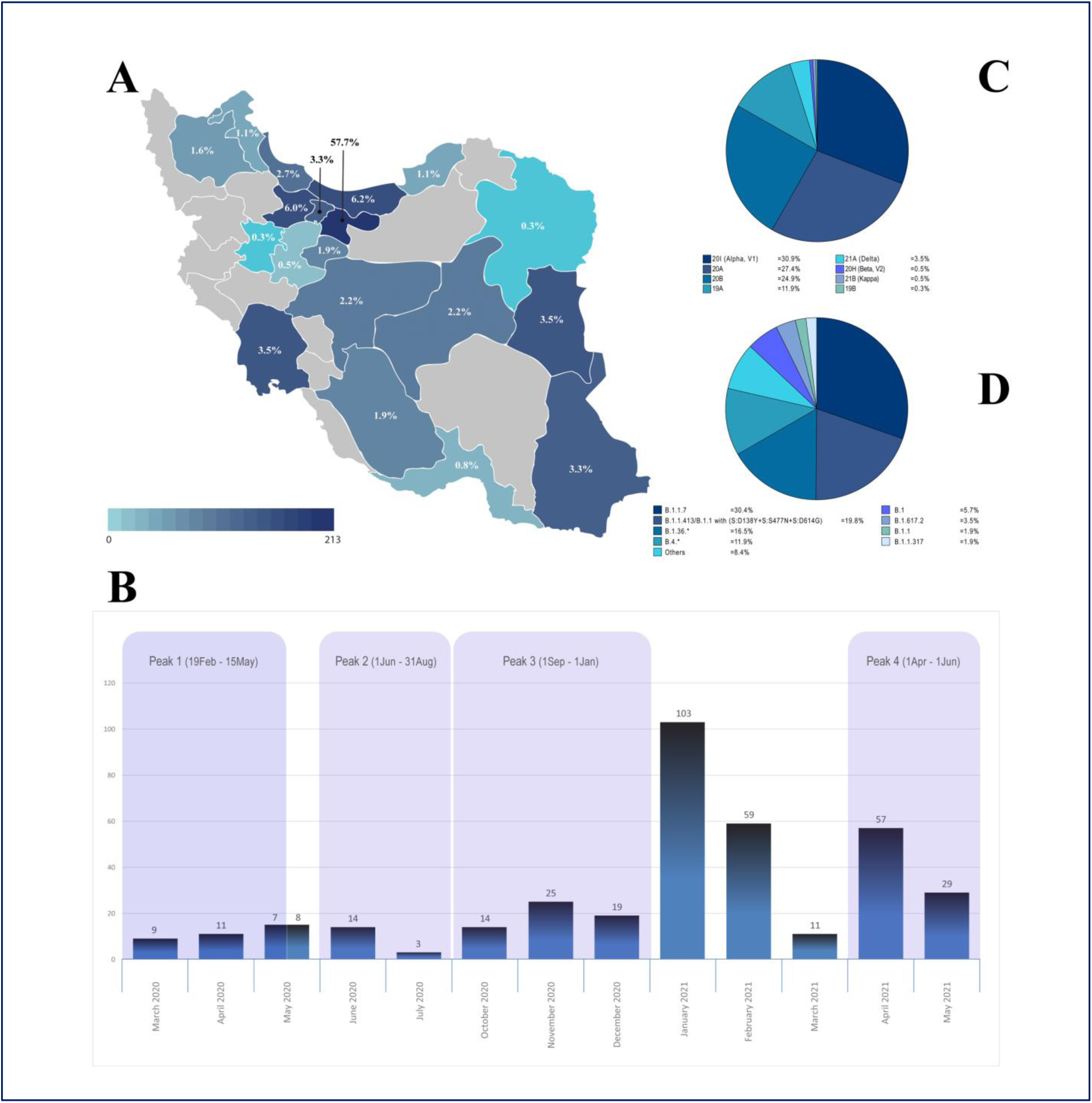
Distribution of SARS-CoV-2 genome sequences in this study; Geographical (A) and chronological (B) and major circulating SARS-CoV-2 clades (C) and lineages (D) in Iran.

### Major circulating SARS-CoV-2 Lineages and Clades in Iran

Over one year of SARS-CoV-2 epidemic, we detected at least 35 different lineages and eight clades circulating in the country. The most frequent clades assigned by Nextclade are 20I (Alpha, V1), 20A, 20B, and 19A with the frequency of 31%, 27%, 25%, and 12%, respectively (Fig.1C). The 35 lineages comprise ∼2.7% of Pango lineages circulating the world (O’Toole et al. 2021), with B.1.1.7, B.1.1.413/B.1.1 carrying (S:D138Y+S:S477N+S:D614G), B.1.36.* and B.4.* being the most frequent lineages observed, also representing each of the disease waves in the country, with the frequency of 30%, 20%, 16.5%, and 12%, respectively (Fig.1D).

### Monthly investigation of prominent clades and lineages of SARS-CoV-2 epidemic in Iran

The epidemic in Iran started with 19A clade/B4 lineage (Fig.2-3), imported from China, which dominated the first outbreak peak and existed significantly untill the end of June 2020 (Fattahi et al. 2021). In May 2020, the 20A clade was observed in our cohort for the first time, represented by B.1 and B.1.36 lineages. The B.1.36 is more prominent and shows an increasing ratio from June 2020 to the end of February 2021. The 14 samples submitted by other centers in GISAID in July-September 2020 (TableS2), also confirmed the similar pattern of the increase in 20A clade; mostly B.1.36 lineages (Not shown in Fig.2, TableS2). Therefore, the 20A clade (leading by B.1.36 lineage) can be considered as one of the causes of the second disease wave in summer 2020, possibly along with B.4 (Fig.3). The 20B clade was first detected in June 2020 (B.1.1.317: Russian lineage), but the prominence of this clade can be observed from October 2020, leading by a distinct B.1.1 lineage carrying the three D138Y, S477N, and D614G mutations on the spike. This lineage constitutes ∼80% of the 20B clade, prompting the third disease wave in autumn 2020 in cooperation with the 20A clade (leading by B.1.36 lineage) (See Fig.2-3. and Fig.S1 for a detailed overview).

**Figure 2.**
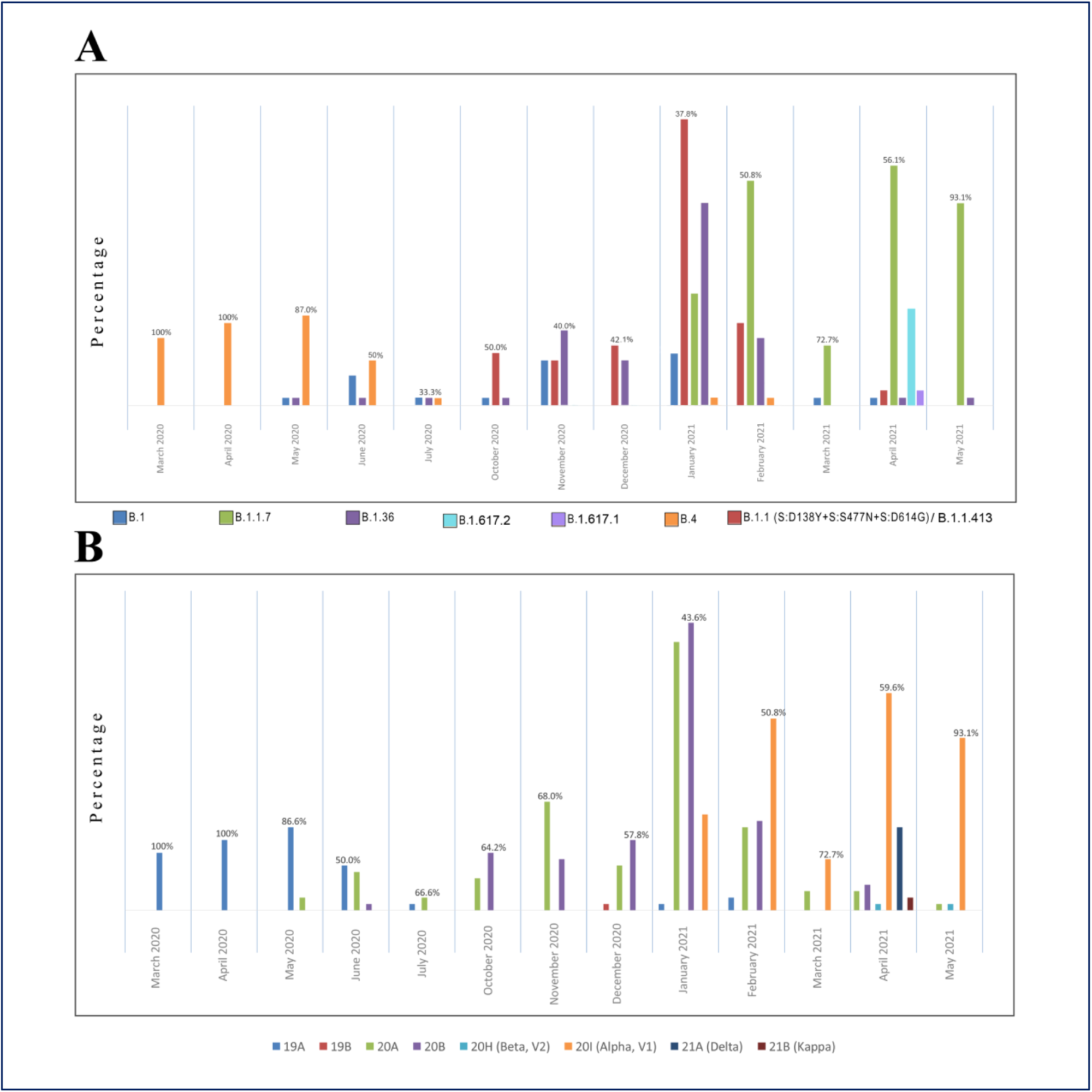
Monthly investigation of prominent lineages (A) and clades (B) of SARS-CoV-2 epidemic in Iran.

**Figure 3.**
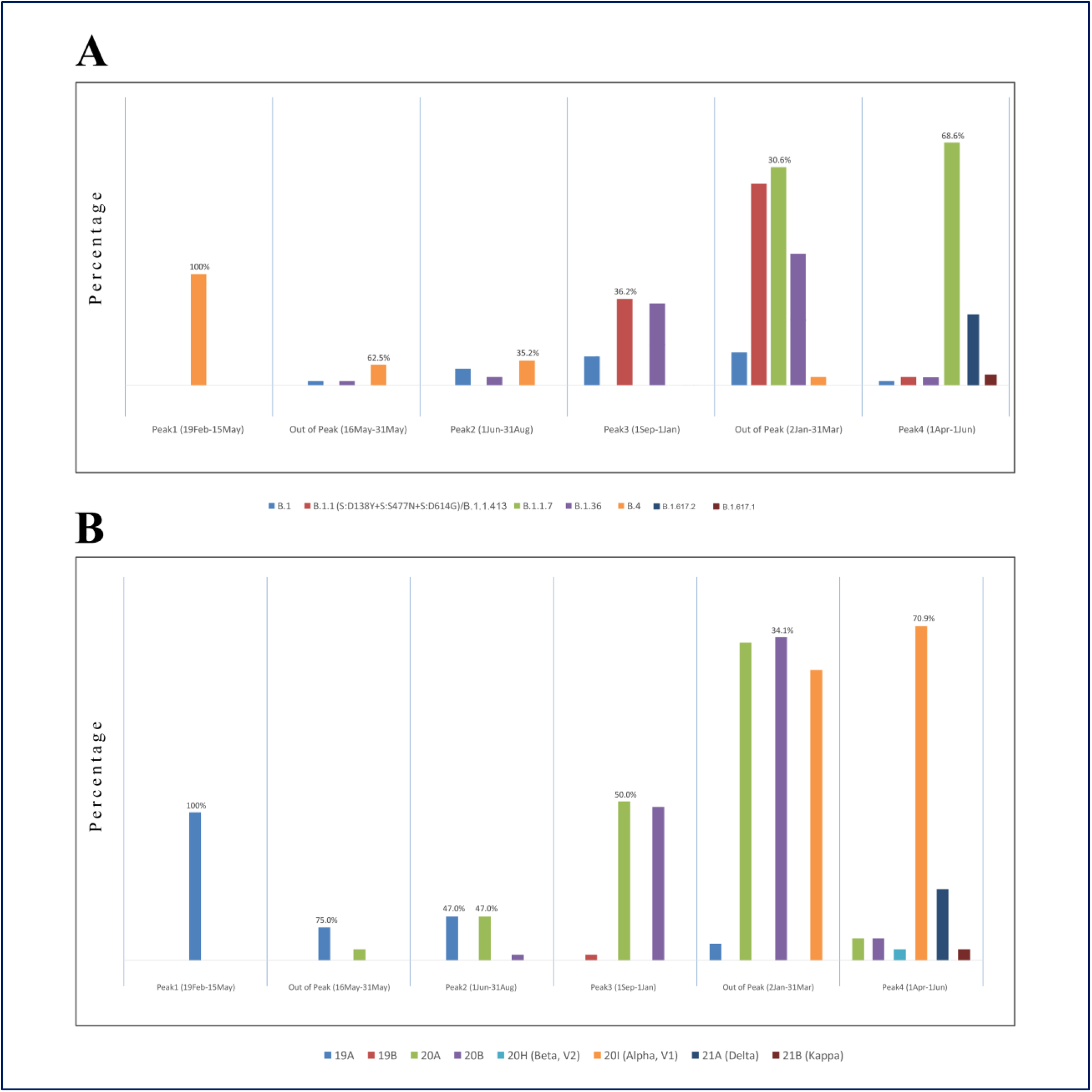
Prominent lineages (A) and clades (B) based on the disease waves of SARS-CoV-2 epidemic in Iran.

The specific mutational combination of [D138Y-S477N-D614G] in spike is initially observed in sequences of October 2020, in which all are assigned as B.1.1 (carrying R203K/G204R) by Pangolin v3.1.7. We further explored the presence of such a spike mutational pattern in the world and detected those in sequences from Brazil (some assigned as B.1.128), UK, USA, Russia (assigned as B.1.1.317 with S:D138Y+S:S477N+S:A845S combination) (Klink et al. 2021). Finally, upon the increase in the number of similar sequences in GISAID and mostly from this cohort, ∼ 80% of these distinct sequences could be finally assigned as B.1.1.413 by Pangolin v.3.1.11 (2021-08-24). The B.1.1.413 carries the following characterizing mutations of [ORF1a:S944L (nsp3: S126L), ORF1b:G128C (RdRp:G137C), ORF1b:P314L (RdRp:P323L), S:D138Y, S:S477N, S:D614G, M:I73M, ORF8:G66E, ORF8:del67/68, N:R203K/G204R] (Latif et al. Accessed 8 September 2021.). It seems that B.1.1.413 is significantly prominent in the SARS-CoV-2 epidemic in Iran. However, this lineage was initially detected on 7 September 2020 in the UK (EPI_ISL_566603), and at least four other similar sequences from Australia and Canada are available in GISAID before its first detection in Iran (24 October 2020), proposing the possible import of this lineage rather than being formed in the country. Still, we cannot accurately determine its earliest date in Iran because of a gap in sequences of August-September 2020. Overall, we introduce B.1.1.413, as a characterizing lineage in Iran in autumn-winter 2020 infection wave.

The abovementioned clades dominated the epidemic untill the end of January 2021, when the first 20I (Alpha, V1) clade/B.1.1.7 lineage was detected in Tehran. Subsequently, the increase in the 20I clade can be observed until May, constituting 93% of the sequences and mediating the 4^th^ disease wave in spring 2021. However, as shown in Fig.2, the 20A and 20B did not entirely disappear and still could be detected in random samples but with a diminishing ratio until May. Due to the non-random sequencing of the suspected samples obtained through prior spike region screening of a larger volume of viral isolates, the contribution of B.1.1.7 sequences in preceding months to the fourth outbreak peak is slightly skewed. But still, a growing slope is noticeable until April-May 2021, when almost all the random samples belonged to B.1.1.7 lineage. Similarly, the surge of Delta variant in April belongs to the non-random samples and does not show a high frequency of this variant at that point.

### Diversity and distribution of Variants of Concern (VOCs) and Variants of Interest (VOIs) in SARS-CoV-2 epidemic in Iran

Overall, the Alpha, Beta, and Delta variants were distinguished in SARS-CoV-2 samples of our cohort. The Alpha variant (B.1.1.7) comprised 31% of our cohort corresponding to the 4^th^ disease wave. The first sequence of B.1.1.7 was obtained in January 2021 through screening of passengers in Tehran. Tracking the viral isolates in the upcoming months showed a fast-growing replication; starting from 14.6% of samples in January, passing by more than 50% of samples in March, and reaching almost 93% of random samples in May. The Beta variant (B.1.351) was detected in two non-random samples from the south (Hormozgan province) and southeast of Iran (Sistan and Baluchestan province) in April-May 2021 but did not cause a separate outbreak peak.

The Delta variant (B.1.617.2) was first detected in non-random samples from Yazd and Qom provinces collected in April 2021 and then in a random sample in the same month, alarming that this variant was already dispersed in the Capital, which eventually led to the 5^th^ disease wave in summer 2021. The dominancy of the Delta variant in this peak is confirmed by Sanger sequencing screening of the spike region of random samples from Tehran (Unpublished data). The Kappa variant (B.1.617.1) was the only VOI detected in non-random samples from Alborz province in April and there is a possibility that this variant is accompanying the Delta variant in the formation of the 5^th^ outbreak peak.

Furthermore, A.23.1 (Variant of Note) was once detected in our cohort; a sub-lineage of A.23, dominating the Uganda epidemic in March 2021 (Bugembe et al. 2021) and the only A lineage (19B clade) observed in the SARS-CoV-2 epidemic in Iran. The A.23.1 is now an international lineage carrying F157L, V367F, Q613H, and P681R mutations of potential biological concern. This lineage was detected in a random sample from Tehran in December 2020 but was not seen any longer in the upcoming months.

### Mutation surveillance of SARS-CoV-2 epidemic in Iran

The 31 proteins encoded by 14 open reading frames (ORFs) in the SARS-CoV-2 genome are arranged into 16 non-structural proteins (Nsps; encoded by ORF1ab gene region), 4 structural proteins (Spike; S, Envelope; E, Membrane; M, and Nucleocapsid; N) and 11 accessory proteins (ORF3a, ORF3b, ORF3c, ORF3d, ORF6, ORF7a, ORF7b, ORF8, ORF9b, ORF9c, and ORF10) (Redondo et al. 2021). Although Coronaviruses have shown higher fidelity in their replication, still thousands of spontaneous mutations have been accumulated in the SARS-CoV-2 genome, since its emergence in 2019, and 79% of its amino acids are mutated at least once (Jaroszewski et al. 2020). Mutation investigation in SARS-CoV-2 genomes provides reliable information about the epidemic status, as these mutations may have effects on viral transmission rate in addition to the host’s antiviral immune response (Leary et al. 2021).

In this cohort, 1577 distinct nucleotide mutations were identified in total, of which 853 (54%) disturb SARS-CoV-2 proteome; including 825 (96.6%) missense, 22 (2.6%) deletion and 6 (0.7%) Stop gain/loss mutations. Of the 1577 distinct nucleotide mutations, 1068 (67.7%) were observed only once in the sequences (1050 substitutions and 18 deletions). Similar to the other studies (Jaroszewski et al. 2020), missense mutations comprised the largest percentage of nucleotide mutations (52.5%).

### Top mutations and the frequent co-occurrences of mutations

In this study, the ten top frequent mutations are explored regardless of the sequence collection dates (Table1). Among these, six mutations represent >50% of the sequences; including four missense mutations (D614G, P323L, R203K, and G204R), one synonymous (3037C>T; F105F), and one non-coding variant (241C>T). Therefore, the top mutation list of the SARS-CoV-2 epidemic in Iran is similar to the other parts of the world, as 88.6% (317/358) of the sequences in Iran carry the quadruplet of [A23403G-C14408T-C241T-C3037T] which are the most frequent mutations worldwide (Wang et al. 2021), corresponding to the G clade (Mercatelli and Giorgi 2020). This quadruplet of mutations co-occurs with R203K/G204R in 53% (188/358) of sequences in Iran; the two subsequent top mutations defining the GR clade (Mercatelli and Giorgi 2020). Furthermore, the Q57H mutation, which is the third top missense mutation in the USA (Wang et al. 2021), is observed in 28% (99/358) of our cohort but it’s not among the ten top list. This reflects the similarity of the Iranian SARS-CoV-2 epidemic more into Europe rather than North America.

**Table 1.**
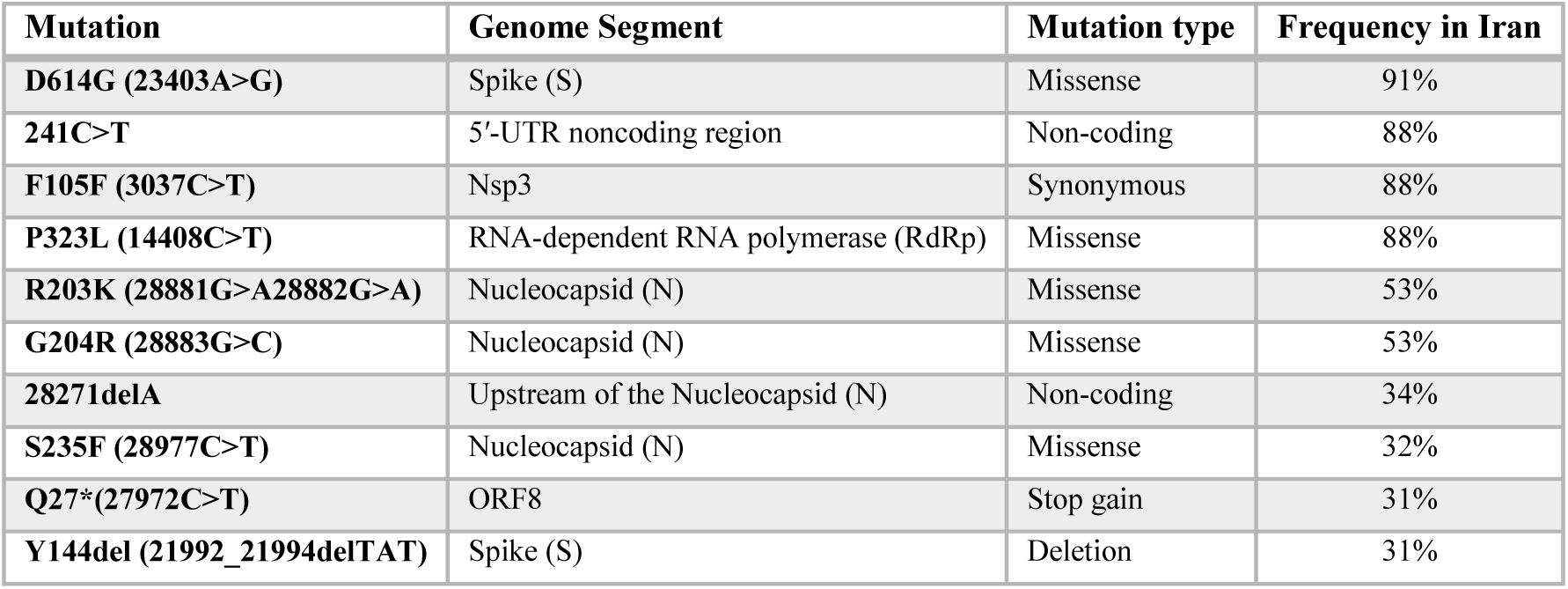
The 10 top frequent mutations in SARS-CoV-2 epidemic in Iran

| Mutation | Genome Segment | Mutation type | Frequency in Iran |
| --- | --- | --- | --- |
| **D614G (23403A>G)** | Spike (S) | Missense | 91% |
| **241C>T** | 5'-UTR noncoding region | Non-coding | 88% |
| **F105F (3037C>T)** | Nsp3 | Synonymous | 88% |
| **P323L (14408C>T)** | RNA-dependent RNA polymerase (RdRp) | Missense | 88% |
| **R203K (28881G>A28882G>A)** | Nucleocapsid (N) | Missense | 53% |
| **G204R (28883G>C)** | Nucleocapsid (N) | Missense | 53% |
| **28271delA** | Upstream of the Nucleocapsid (N) | Non-coding | 34% |
| **S235F (28977C>T)** | Nucleocapsid (N) | Missense | 32% |
| **Q27\*(27972C>T)** | ORF8 | Stop gain | 31% |
| **Y144del (21992_21994delTAT)** | Spike (S) | Deletion | 31% |

The rest of the sequences (11%) carry [G1397A-T28688C-G29742T] mutations of B.4 lineage. While dominant (>70% of samples) at the early phase, now, passing more than one year from the start of the epidemic, these mutations are disappeared and replaced by [C241T-C3037T-C14408T-A23403G] upon the entry of the B.1.* lineages in late February 2020 (Fattahi et al. 2021). Moreover, the most frequent missense mutation at the early phase was V198I located in Nsp2, which was then replaced by the renowned D614G mutation (91%). D614G was first detected on 1 May 2020 in Iran and not typically along with [C241T-C3037T-C14408T] but with the defining mutations of B.4 lineage [G1397A-T28688C-G29742T]. Later in the same month (18 May 2020), we could detect this mutation accompanying [C241T-C3037T-C14408T], which also carries the second top missense mutation in our cohort; P323L. We conclude that all viral isolates in Iran except one assigned as the A23.1, belong to the B lineage, carrying one of the abovementioned co-occurrence of mutations. Remarkably, no rise based on the entry of “A lineage” was observed in the country.

One of the other top 10 mutations is 28271delA (34%), a non-coding deletion upstream of the Nucleocapsid gene that entered the country by B.1.1.7 lineage. This deletion interacts with other defining mutations of B.1.1.7, such as N501Y, P681H, and T716I (Yang et al. 2021), and along with N:D3L (28280G>C,28281A>T,28282T>A), which is also frequently observed (30%) in our cohort, can cause the high viral transmissibility of B.1.1.7 lineage. The next top mutations; S235F, Q27*, and Y144del, are also among the defining B.1.1.7 mutations, and minor differences in their frequency can be explained by the variability in sequence qualities which can affect the subsequent mutation detection.

In the following, we investigated our cohort for the fifteen most frequent missense variants (>5%) in the world, reported in Miao et al. study. As shown in Table2, we shared the top 4 missense variants with the world. The Q57H, S194L, S477N, and L37F mutations are present with high frequencies in Iran but are not among the fifteen tops. Besides, some others (A222V, V30L, A220V, T85I, L18F, S24L, I120F) have no or considerably low frequency and occur typically in 20C, 20E, 20F, and 20G clades which are absent from the SARS-CoV-2 epidemic in Iran (Table2). On the other hand, the subsequent 11 top missense mutations in Iran are mostly the defining B.1.1.7 mutations. The observed difference is largely caused by the different time intervals of the two cohorts rather than the distinct genetic diversity of SARS-CoV-2 in Iran compared to the world. Miao et al. investigated 260,673 sequences between December 2019-12 January 2021, before the dominancy of B.1.1.7 lineage in the world (Miao et al. 2021).

**Table 2.** Investigation of the world fifteen top mutations (until January 2021) to SARS-CoV-2 epidemic in Iran

| Variants | SARS-CoV-2 proteome | Frequency in world | Frequency in Iran | Iran/World | Main substitutions characterizing the SARS-CoV-2 clades |
| --- | --- | --- | --- | --- | --- |
| D614G | Spike | 93.88% | 91% | Top 1/ Top 1 | 20A, 20A.EU2, 20B, 20C, 20E (EU1), 20H, 20I |
| P323L | NSP12 | 93.74% | 88% | Top 2/ Top 2 | 20A, 20A.EU2, 20B, 20C, 20E (EU1), 20H, 20I |
| R203K | Nucleocapsid | 28.45% | 53% | Top 3/ Top 3 | 20B, 20I/501Y.V1 |
| G204R | Nucleocapsid | 28.13% | 53% | Top 4/ Top 4 | 20B, 20I/501Y.V1 |
| A222V | Spike | 26.14% | 2% | Not among the 15 Top variants/ Top5 | 20E (EU1) |
| V30L | ORF10 | 26.01% | Not detected | Not among the 15 top variants / Top6 | 20E (EU1) |
| A220V | Nucleocapsid | 25.96% | 2% | Not among the 15 top variants / Top7 | 20E (EU1) |
| Q57H | ORF3a | 23.60% | 28% | Not among the 15 top variants / Top8 | 20C, 20H/501Y.V2 |
| T85I | NSP2 | 15.38% | 0.6% | Not among the 15 top variants / Top9 | 20C, 20H/501Y.V2 |
| L18F | Spike | 12.08% | 0.3% | Not among the 15 top variants / Top10 | NA |
| S194L | Nucleocapsid | 6.29% | 22% | Not among the 15 top variants / Top11 | NA |
| S477N | Spike | 6.62% | 22% | Not among the 15 top variants / Top12 | 20A.EU2, 20F |
| L37F | NSP6 | 6.52% | 14% | Not among the 15 top variants / Top13 | 20G |
| S24L | ORF8 | 5.24% | Not detected | Not among the 15 top variants / Top14 | 20G |
| I120F | NSP2 | 5.23% | Not detected | Not among the 15 top variants / Top15 | 20F |

### Temporal Trend of SARS-CoV-2 genome mutations in Iran

To provide a comprehensive picture of SARS-CoV-2 genomic diversity in Iran, we analyzed variations of the SARS-CoV-2 proteome in a monthly manner. The average number of mutations per sequence in our cohort is 20. We observed two different tendencies in the mean number of mutations ranging from 14.9 before the entry of B.1.1.7 lineage, then rising into 34.4 from January 2021, which is rational as this lineage introduces 23 new mutations into the SARS-CoV-2 genome sequence (Rambaut et al. 2020). Then, we monitored the emergence of mutations and the distribution of top frequent mutations (>20%) in each month and each disease wave of SARS-CoV-2 epidemic in Iran (Fig.4, Fig.S2-S6).

**Figure 4.**
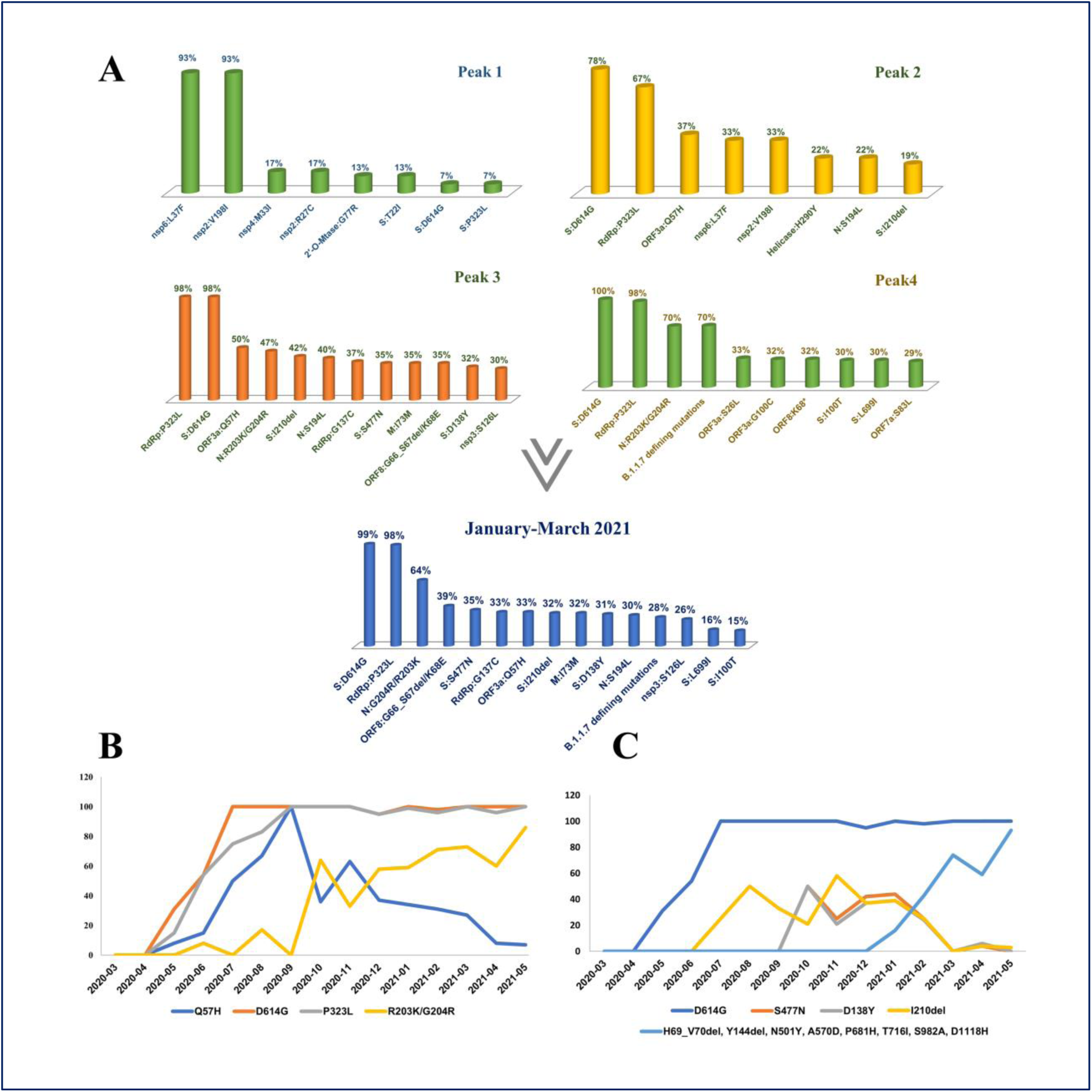
Top mutations based on the disease waves of SARS-CoV-2 epidemic in Iran (A). Chronological trend of top lineage-defining (B) and Spike (C) mutations.

### Top mutations in the first wave of SARS-CoV-2 infection

The top missense mutations commonly dominating the first outbreak peak (March-May 2020) are nsp6:L37F and nsp2:V198I, while only constituting 14% and 11% of the total mutations, respectively (Fig.4A, Fig.S2). The frequency of these two mutations decreased gradually; as in August 2020, they were not among the top 20% mutations and then completely disappeared from the epidemic along with the B4 lineage, comparable to the decaying global tendency reported for L37F.

The nsp6:L37F mutation has shown a significant correlation to asymptomatic SARS-CoV-2 infections and is unfavorable for viral transmission, possibly by compromising the ability of the virus to confront the innate cellular defense (Wang et al. 2020). This may partly explain the unrecognized transmission of the disease at the start of the epidemic in Iran. However, misdiagnosed patients or limited testing capacities are among the other reasons (Ghafari et al. 2020, Fattahi et al. 2021).

The first and prominent spike mutation in the first disease wave was T22I (13%), which was then replaced upon the entry of the D614G mutation in May 2020. Overall, the frequency of T22I mutation in our cohort is considerably low (2%) and disappeared from July 2020 onwards. However, this mutation is still circulating in the world, as of August 2021 (Latif et al. Accessed 5 September 2021). Moreover, the top frequent S:D614G and RdRp:P323L mutations in our cohort started to appear from the end of May 2020 and gradually increased in the following months, raising the second disease wave (Fig.S2).

### Top mutations in the second wave of SARS-CoV-2 infection

The top missense mutations commonly dominating the second outbreak peak (June-August 2020) are S:D614G, RdRp:P323L, and ORF3a:Q57H, while constituting 91%, 88%, and 28% of the total mutations, respectively (Fig.4A). This new mutational tendency is correlated with the entry of B.1* lineages and dominancy of 20A clade in the second peak and is comparable to the world as the D614G mutation gradually dominated the pandemic while mostly accompanies P323L mutation (Vilar and Isom 2021).

The renowned D614G mutation is located outside the Receptor Binding Domain (RBD) of spike protein but possibly increases the viral transmission. Additionally, the P323L mutation encodes a more error-prone RdRp, correlated with an increase in viral genetic diversity, and finally allows the virus to spread more effectively in different environmental conditions and populations (Banoun 2021). Therefore, the appearance of the second outbreak peak can be justified by such a new mutational trend in the country.

The next prominent mutation in this peak is ORF3a:Q57H, first detected in May 2020, gradually increased in the second disease wave, also sustained in the third peak (50%), and then disappeared from the top 20% mutations of the fourth peak (Fig.4B, Fig.S2-6). ORF3a is an essential protein for viral cytotoxicity and is known to activate the inflammasome. This mutation causes a dramatic change in protein structure, affecting the binding affinity of Orf3a–S and Orf3a–Orf8 protein interactions, and finally may be correlated with enhanced viral virulence (Banoun 2021, Wu et al. 2021). This dramatic change in protein structure is also reported for S194L and R203K/G204R in Nucleocapsid. The S194L was first detected in July 2020, reaching a frequency of >20% in the second peak, and shows a similar mutational tendency as Q57H. Furthermore, as the B4 lineage was still circulating in the country, we can still observe the nsp6:L37F and nsp2:V198I mutations (33%), but with significantly decreased frequency compared to the first peak (Fig.4A, Fig.S3)

The prominent spike mutation other than D614G in this peak is I210del, first observed in July 2020, and holding a similar mutational tendency as Q57H and S194L (Fig.4C). This deletion is mainly detected in B.1.36* sequences (73%) in Iran. However, its co-occurrence with S:D138Y, S:S477N, and also B.1.1.7 sequences is seen but never led to a significant rise in the frequency.

### Top mutations in the third wave of SARS-CoV-2 infection

The top missense mutations commonly dominating the third outbreak peak (September-December 2020) are S:D614G, RdRp:P323L, N:R203K/G204R, and ORF3a:Q57H, while constituting 91%, 88%, 53%, and 28% of the total mutations, respectively (Fig.4A, Fig.S4). This new mutational tendency is correlated with the prominence of 20B clade since October 2020, leading by B.1.1.413.

The B.1.1 lineage is designated by the three adjacent nucleotide substitutions; 28881G>A, 28882G>A, and 28883G>C. These arose on the background of D614G mutation through homologous recombination and code adjacent R203K/G204R in Nucleocapsid (Mercatelli and Giorgi 2020, Leary et al. 2021). The R203/G204 amino acids are located in the serine/arginine-rich (SR-rich) linker region of Nucleocapsid, which is involved in modulating Nucleocapsid multimerization, and cellular localization. Therefore, R203K/G204R mutations may destabilize the protein, followed by a decrease in the overall structural flexibility, and finally can increase the virulence of SARS-CoV-2 (Leary et al. 2021, Rahman et al. 2021, Troyano-Hernáez et al. 2021, Wu et al. 2021)

Consequently, the accumulation of such mutations to the mutational burden of the epidemic can explain the third disease wave in autumn 2020. Besides, RdRp:G137C and M:I73M mutations also show high frequency in this peak, as these are characteristic mutations of B.1.1.413 lineage.

The prominent spike mutations other than D614G in the third disease wave are I210del, S477N, and D138Y. The co-occurrence of D138Y, S477N, and D614G is commonly observed in B.1.1.* lineages (84%) of our cohort and their prominence are the characterizing point of this wave of infection.

D138Y is one of the defining mutations of P.1, a sublineage of B.1.1.28. It is located in the N-terminal domain (NTD) of the S1 protein, and accompanying other P.1 defining mutations in the NTD reduces neutralization of mAb159 by disrupting its epitope (Dejnirattisai et al. 2021). However, D138Y mutation in our cohort does not accompany other P.1 mutations in NTD, such as L18F, T20N, P26S, and R190S, but co-occurs with S477N and D614G. As a result, we cannot justify whether the D138Y accompanying other spike mutations (S477N-D614G) still shows the reduction in mAb159 neutralization.

However, this combination itself is remarkable. S477N is the most frequent RBD mutation that often co-occurs with D614G (Singh et al. 2021). S477N increases viral affinity for ACE2 receptor and is known to escape multiple mAbs, although this neutralization resistance was not observed in convalescent serum (Harvey et al. 2021, Singh et al. 2021). The S477N mutation has arisen multiple times independently. It was observed with a global frequency of 4-7% in mid-June 2020. Then, it dominated the majority of European sequences in autumn 2020 (20A.EU2), also the winter outbreak in Oceania (Harvey et al. 2021, Hodcroft et al. 2021). The current study shows that S477N was first detected in October 2020 in Iran and was one of the frequent spike mutations not only in autumn 2020 but in winter 2021 (Fig.4C). Later, the S477N and D138Y were seen accompanying some B.1.1.7 sequences but never showed a significant rise.

### Top mutations in the fourth wave of SARS-CoV-2 infection

In this study, the time interval between the official third and fourth peaks (January-March 2021) was also carefully investigated, based on the emergence of B.1.1.7 VOC in the UK. We aimed to track the entry and accumulation of this VOC, also chasing the fate of circulating lineages in autumn 2020. The B.1.1.7 was first detected in September but notified as an outbreak in December 2020 in the UK. Similarly, after its first detection in January 2021 in Iran and with a 3-month gap, the surge in hospitalization and deaths was officially observed from April 2021. However, in some southern provinces such as Khuzestan, the corresponding disease wave was observed sooner in winter 2021.

The top missense mutations commonly dominating this time interval are S:D614G, RdRp:P323L, and N:R203K/G204R, the same as the last peak. However, the frequency of Q57H started to drop by, while we see an increasing slope for N:R203K/G204R (Fig.4B).

The observed mutational trend is correlated with the increase in 20I and decrease in 20A and 20B clades from January to March 2021. Therefore, according to our expectation, and based on the approximate parallel frequency of 20A, 20B, and 20I clades in these months, the prominent spike mutations other than D614G are S477N, I1210del, D138Y, and the defining B.1.1.7 spike mutations; H69_V70del, Y144del, N501Y, A570D, P681H, T716I, S982A, and D1118H (Fig.4A, Fig.S5).

Afterward, the seventeen B.1.1.7 defining mutations are added to the list of top mutations (70%) in the fourth outbreak peak (April-May 2021) (Fig.4A, Fig.S6). Furthermore, two new spike mutations are observed in this disease wave, I100T and L699I, with a frequency of 30%. Indeed, half of the B.1.1.7 sequences (46.5%) carry the defining B.1.1.7 spike mutations in addition to these two, while the other half (53.5%) carried the defining mutations uniquely or accompanying other random spike mutations. These mutations were both detected from January, the time of B.1.1.7 entry to the country, which proposes being imported rather than created during the replication of the original variant. This combination gives a low global frequency of 0.5%, while Iran shows the most cumulative prevalence (Latif et al. Accessed 7 September 2021., Latif et al. Accessed 7 September 2021.). Therefore, we conclude the involvement of these two mutations in B.1.1.7 sequences as a characteristic feature of the B.1.1.7 epidemic in Iran. However, their contribution to the epidemic drops by a third compared to the typical B.1.1.7 sequences (Fig.S6).

In April 2021, a non-random increase in the frequency of B.1.617.2 VOC (24%) explains the lower ratio of B.1.1.7 mutations in this month compared to March (Fig.S5-6). Furthermore, G142D spike mutation can be observed in 75% of B.1.617.2 sequences, in addition to its typical defining mutations (Fig.S6). However, this is not a specific observation, as G142D accompanies 64% of B.1617.2 sequences worldwide (Latif et al. Accessed 11 September 2021.).

### Mutational profile of SARS-CoV-2 viral components in Iran

We also investigated the most recurrent mutations in each SARS-CoV-2 protein (Fig.5A), as these proteins have shown different mutation rates, correlated with their structural and functional features (Jaroszewski et al. 2020, Vilar and Isom 2021). The majority of mutations disturbing coding sequences in our cohort occurred in the ORF1ab gene region (58%), which is expected as it covers 71% of the SARS-CoV-2 genome, followed by the spike (17%) and Nucleocapsid (8%) genes; being also the second and third-longest genes (Fig.5B). However, this does not reflect the higher mutation rate as the largest number of distinct mutations in these genes can be partly due to their largest genomic length. So, when the distinct occurrence of mutations in each gene is adjusted to its length (Fig.5C), the pattern changes and a higher mutation rate can be observed in accessory proteins (ORF8, ORF7b, ORF3a, and ORF7a) and structural proteins such as Nucleocapsid and Spike, rather than the non-structural genes such as nsp12 (RdRP). This follows the previous information considering the ORF1ab region as under-mutated compared to the structural and accessory proteins. Nsp proteins and especially nsp12 (RdRP) have pivotal roles in RNA transcription and viral replication and therefore should be more intact. Whereas structural and accessory proteins such as ORF8 and N proteins are involved in virus-host interactions and the over-mutation is adjusting the virus virulence or host immune avoidance (Jaroszewski et al. 2020, Vilar and Isom 2021). Consistent with this, the recurrent mutations of the following genes showed a frequency of less than 10% in our cohort (Red circles in Fig. 5A); 11 Nsp genes (nsp1, nsp4, nsp5, nsp7, nsp8, nsp9, nsp10, nsp13, nsp14, nsp15, nsp16), one structural protein (E) and three accessory proteins (ORF6, ORF7b, and ORF10).

**Figure 5.**
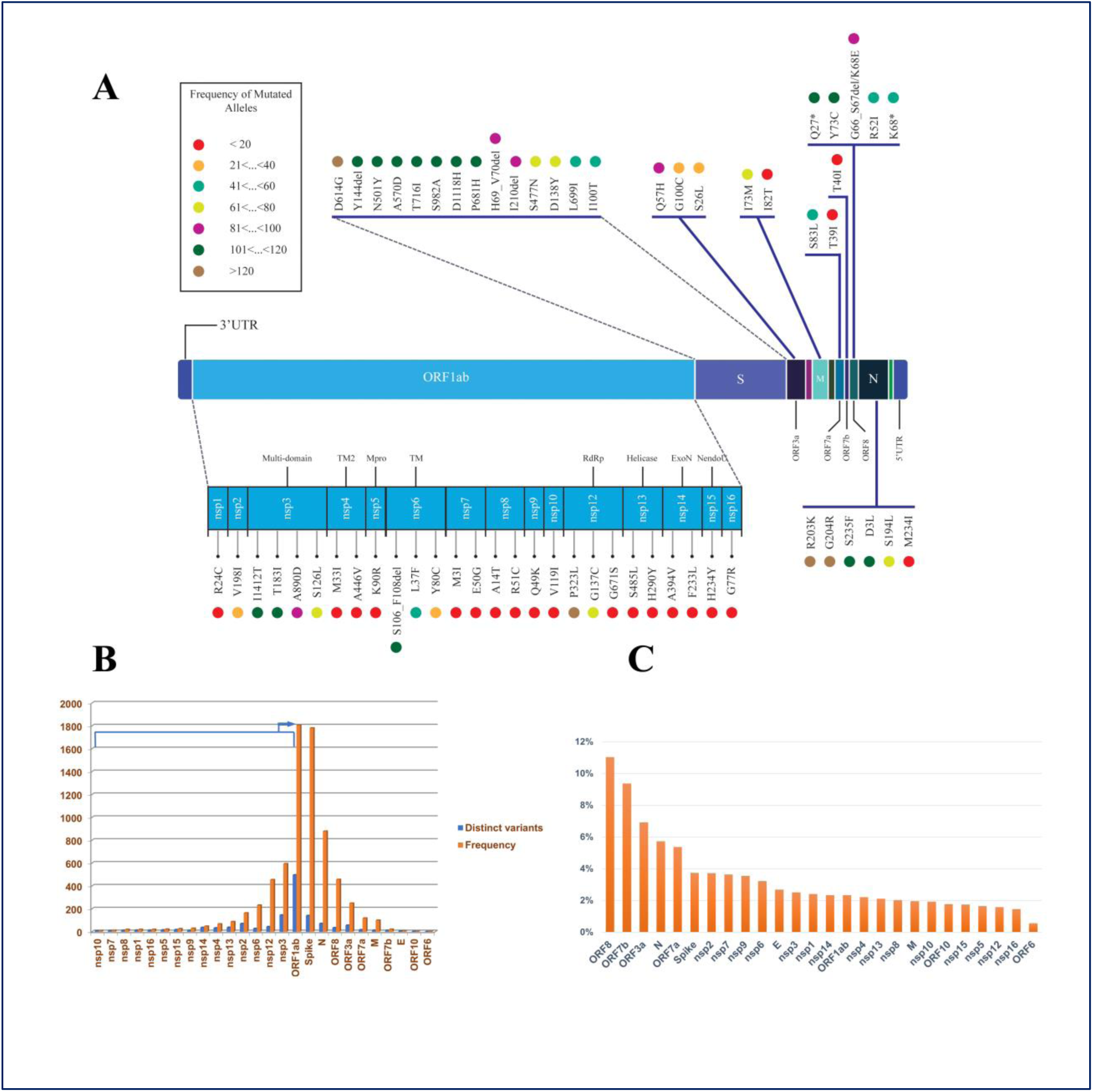
Mutational profile of SARS-CoV-2 viral components in Iran; **(A)** shows a schematic representation of SARS-CoV-2 genes. The prominent mutations of each gene are drawn in colorful circles. The colors are based on the mutation frequencies indicated in the left panel; **(B)** shows the mutation rate of SARS-CoV-2 genes based on the number of distinct mutations (Blue) and frequency of the mutations (Orange) in each gene, **(C)** shows the adjusted mutation rate of each SARS-CoV-2 gene based on its length.

Among the structural proteins, Envelope (E) shows the lowest number of mutations with literally no recurrent mutation in our cohort (only 6 missense variants occurred once). The Envelope is a small protein constituting the virus protein interface with the external environment along with Spike and Membrane (Bianchi et al. 2020). This protein is involved in critical aspects of the viral life cycle, such as assembly, release, and virulence phases (Schoeman and Fielding 2019, Chai et al. 2021), and has shown the lowest mutation rate among the other SARS-CoV2 structural proteins (Vilar and Isom 2021).

Among the non-structural proteins comprising the ORF1ab proteins, nsp3, nsp2, and nsp12 (RdRP) showed the largest number of mutations in our cohort, with the percentages of 17%, 8%, and 5%, respectively (Fig.5B). As expected, the nsp12 (RdRp) re-locates into the lowest limit of mutation rate, when the number of mutations is adjusted to its length (Fig.5C).

Of nsp12 (RdRp) recurrent mutations, the top (20%) ORF1b:P314L (14408C>T; RdRp:P323L) and ORF1b:G128C (13849G>T; RdRp:G137C) are not located in the RdRp-RNA interface, which is the most under-mutated region of this gene. On the other hand, the nsp3 recurrent mutations are nsp3:I1412T (6954T>C; ORF1a:I2230T), nsp3:T183I (3267C>T; ORF1a:T1001I), nsp3:A890D (5388C>A;

ORF1a:A1708D), and nsp3:S126L (3096C>T;ORF1a:S944L). Although nsp3 protein is among the over-mutated SARS-CoV-2 proteins, its C-terminal domain is significantly less mutated. Consistently, the frequent nsp3 mutations in our cohort are outside this domain (Fig. 5B-C) (Jaroszewski et al. 2020).

### Spike mutations in SARS-CoV-2 epidemic in Iran

The Spike protein (particularly its RBD) interacts directly with the ACE2 receptor in human cells and therefore is a preferred target for therapeutic antibodies and vaccines (Min and Sun 2021). Therefore, spike mutations develop major effects on viral infectivity and dissemination of VOCs with global health impacts (Khateeb et al. 2021) and at least, 87% of RBD residues are mutated during the pandemic. However, most of these mutations are not persistent, except those developing VOCs (Li et al. 2021).

In total, 141 spike mutations were detected in the SARS-CoV-2 epidemic in Iran, in which 90% occurred with a frequency less than 5% (TableS3). The most frequent spike mutation is D614G, followed by 13 other frequent mutations based on the prominent lineages circulating in the country in different time intervals; B.1.1.7 (H69_V70del, Y144del, N501Y, A570D, P681H, T716I, S982A, D1118H in addition to I100T and L699I), B.1.1.413 (D138Y, S477N) and B.1.36 (I210del) (Fig. 5A).

Among these, S477N, N501Y, and P681H are suspected to increase viral transmission and virulence or reduce vaccine efficiency. The first two increase the affinity for the ACE2 receptor, while the third acts by locating in the S1/S2 furin cleavage site. The H69_V70del, Y144del, and I210 del are located in NTD; known as recurrent deletion region (RDR), and may have a role in viral evasion from the human immune response. Moreover, D138Y mutation is also located in NTD, a region known to have a substantial role in spike antigenicity (Rambaut et al. 2020, Barton et al. 2021, Harvey et al. 2021, Khateeb et al. 2021, Singh et al. 2021).

In total, 12 different spike mutations are found in RBD (Table.3), of which 10 (83%) are represented with a frequency of less than 5%. All these mutations are previously detected around the world. Furthermore, seven of these mutations are the well-known RBD mutations flagged as mutations of concern and interest (E484K, N501Y, S477N, L452R, T478K, K417N, and E484Q). Interestingly, the S477N prevalence in Iran is higher than its cumulative prevalence in the world, corresponding to the dominancy of characteristic B.1.1.413 lineage. Overall, the spike mutation content of SARS-CoV-2 epidemic in Iran is similar to other parts of the world and no country-specific mutation with a significant raise in the epidemic was detected except the significant rise in combination of [D138Y-S477N-D614G] mutations.

**Table 3.** Spike mutations located in RBD region

| Mutations | Prevalence/<br>Earliest Date<br>in Iran | Type, Region | Prevalence/<br>Earliest Date<br>in World | Corresponding Lineage in Iran |
| --- | --- | --- | --- | --- |
| **N501Y** | 30%, 2020.10.14 | Mutation of interest, RBM | 37%, 2020.02.07 | B.1.1.7 (98), B.1.1.7 + E484K (1), B.1.351 (2), B.1.36.31 (2), B.1.1 (2), B.1.1.413 (1) |
| **S477N** | 22%, 2020.10.24 | Mutation of interest, RBM | 2%, 2020.01.28 | B.1.1.413 (68), B.1.1.7 (6), B.1.1.99 (1), B.1.260 (2), B.1.36.10 (1), B.1.533 (1) |
| **L452R** | 4.5%, 2021.01.19 | Mutation of interest, RBM | 32%, 2020.03.03 | B.1.617.2(13), B.1.617.1 (2), B.1.36 (1) |
| **T478K** | 4%, 2021.04.00 | Mutation of interest (?), RBM | 30%, 2020.03.15 | B.1.617.2(13) |
| **E484K** | 0.8%, 2021.04.03 | Mutation of concern, RBM | 6%, 2020.02.01 | B.1.351(2), B.1.1.7 + E484K (1) |
| **K417N** | 0.6%, 2021.04.03 | Mutation of interest, RBD | 1%, 2020.02.15 | B.1.351(2) |
| **E484Q** | 0.6%, 2021.04.00 | Mutation of interest (?), RBM | < 0.5%, 2020.03.03 | B.1.617.1(2) |
| **F490S** | 0.6%, 2021.01.00 | RBM | < 0.5%, 2020.04.05 | B.1.1.7 (1), B.1.36 (1) |
| **A344S** | 0.3% , 2020.04.13 | RBD | < 0.5%, 2020.03.07 | B.4 (1) |
| **Y365C** | 0.3%, 2021.01.00 | RBD | < 0.5%, 2020.12.29 | B.1.1.7 (1) |
| **P479L** | 0.3%, 2021.04.29 | RBM | < 0.5%, 2020.04.03 | B.1.617.2 (1) |
| **V483A** | 0.3%, 2021.01.00 | RBM | < 0.5%, 2020.02.29 | B.1 (1) |

## Discussion

Today; over one year after the declaration of the COVID-19 pandemic by WHO, genome surveillance projects are frequently performed in all countries with the intention of tracking the spread of SARS-CoV-2 variants (Miao et al. 2021). The current study is the report of complete genome sequencing of 369 SARS-CoV-2 viral isolates of the Iranian outbreak from March 2020 till the end of May 2021.

This study provides a comprehensive picture of temporal and geographical dynamics of the prominent SARS-CoV-2 clades/lineages circulating in Iran and introduces the 19A clade (B.4 lineage) dominating the first disease wave (Spring 2020) (Fattahi et al. 2021), followed by 20A (B.1.36), 20B (B.1.1.413) and 20I (B.1.1.7) clades, dominating second (Summer 2020), third (Autumn 2020) and forth (Spring 2021) disease waves, respectively. It also shows that the COVID-19 patients in winter 2021 were mostly exposed and infected by a mixture of circulating 20A (B.1.36), 20B (B.1.1.413), and 20I (B.1.1.7) clades, competing in a diminishing manner for 20A/20B, paralleled with a growing rise of 20I (B.1.1.7), eventually prompting the fourth outbreak peak in spring 2021. Furthermore, our study provides supporting information about the entry of the Delta variant (21A clade) in April 2021, which gave rise to the 5th disease wave in summer 2021.

This temporal dynamic is comparable to the global perspective; Pandemic was dominated by 19A/19B clades till March 2020 and then replaced by 20A clade (appearance of D614G mutation) till mid-September. However, we never detected any viral isolates from 19B clade (Pango A lineages) in Iran, except one sample from A23.1 lineage (19B clade) in December 2020. The 19B clade did not rise in Iran, while in February 2021 resurgence of this clade was reported in some countries (dominated by A.23.1, A.27 and, A.2.5 lineages) (Murall et al. 2021). From mid-September 2020 onward, the pandemic was dominated by 20E clade, while we detected 20B dominancy, which is more similar to the pandemic in Asia, in which 20B was the leading clade in January (Miao et al. 2021).

Our study highlights three hallmarks of the SARS-CoV-2 outbreak in Iran. First; the primary phase mediated by B.4 lineage. Second; a special raise in a B.1.1 lineage carrying a combination of [D138Y-S477N-D614G] spike mutations in autumn 2020 and winter 2021, recently assigned as B.1.1.413. This lineage can be observed in other regions such as Turkey and Europe. However, Iran seems to be the only country that experienced a disease wave due to the growing prevalence of this lineage at that time. Third; B.1.1.7 outbreak mediated by two types of B.1.1.7 sequences, with 46.5% prevalence of [S:I100T-S:L699I + B.1.1.7 defining mutations] in Iran compared to <0.5% in the world. However, this mutational combination did not show a higher transmission rate compared to the original one.

The sequences obtained through this study provided a good resource for the monthly estimation of the SARS-CoV-2 mutational burden in the country. Continuous monitoring of mutations in the SARS-CoV-2 genome is a requisite in each outbreak, as it facilitates tracking the possible formation of VOCs. Such information can also be used in refining the primers and probes applied in RT-PCR-based diagnosis, that are targeting different genes in the SARS-CoV-2 genome. However, accumulating nucleotide variations can disturb primer binding sites, leading to a high percentage of false-negative test results of even up to 50% in some reports (Alizad-Rahvar et al. 2021). The RdRp and Nucleocapsid genes are among the targets of widely-used commercial kits in diagnostic laboratories in Iran. Our data displayed a high frequency of mutations in the Nucleocapsid gene. It also revealed some recurrent mutations in RdRp with a fixed high frequency, such as P323L, G137C, and recently G671S of the Delta variant. Therefore, the current information could be applied in refining and updating primers, and eventually increasing the quality of PCR diagnostic tests in the country.

This study shows that almost all the previous circulating viral lineages were disappeared with the rise of the Alpha variant, which itself might be replaced upon the entry of the Delta variant in April. Due to accelerated transmission of this new VOC, lack of proper quarantine policies, and slow vaccination rate, the country suffered a dramatic increase in the number of daily deaths, as of September 2021. Although there are still some doubts about the effectiveness of vaccines on the Delta variant, the two widely used vaccines in Iran (ChAdOx1 to-19 (AstraZeneca) and Sinopharm) provide sufficient protection after two doses; 67% for AstraZeneca (Lopez Bernal et al. 2021) and 59% for Sinopharm (“China’s inactivated vaccines effective against Delta variant: study” 2021-08-23), and may efficiently help in outbreak control. In conclusion, the genome surveillance performed in this study provided a comprehensive picture of the SARS-CoV-2 mutation profile in Iran, which is beneficial for the assessment of vaccine and therapeutic efficiency in this population.

## Data Availability

Most of the sequence data provided through this study are already uploaded in the GISAID database (https://www.gisaid.org/) and the remaining is going to be submitted.

## Acknowledgments

Iranian Network for Research in Viral Diseases (INRVD) is acknowledged for help with the sample collection through its collaborative research centers and hospitals. This study was funded by the Iran Vice deputy for Research and Technology at the Iran Ministry of Health and Medical Education, grant number: 99/801/A/6/25942.

