## Supplementary material for "Disease waves of SARS-CoV-2 in Iran closely mirror global pandemic trends": Figure.S1-S6

**FigS1.**The detailed overview of lineages in each month and each outbreak peak.

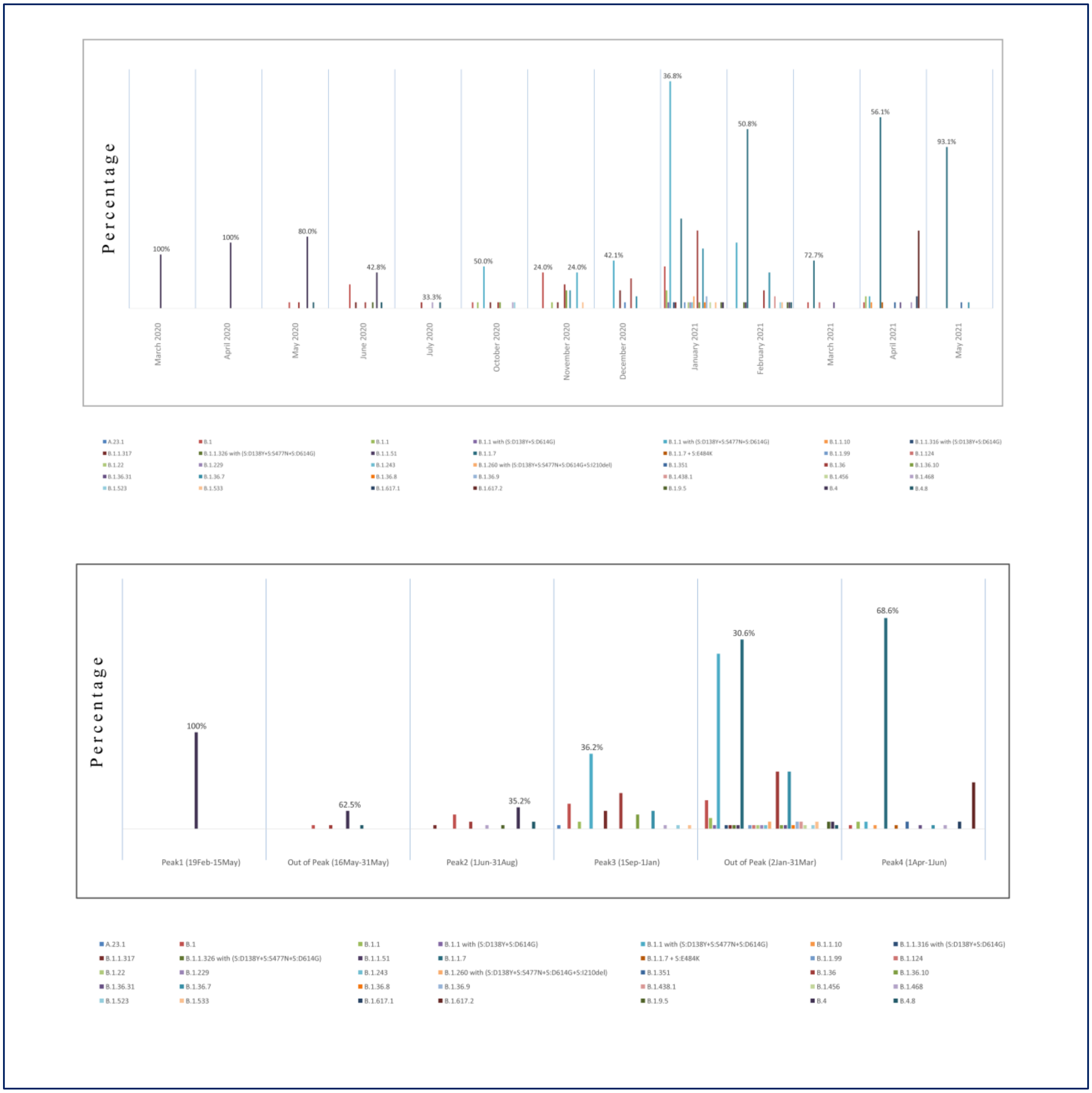

**FigS2.** Top mutations in the first wave of SARS-CoV-2 infection (March-May 2020)

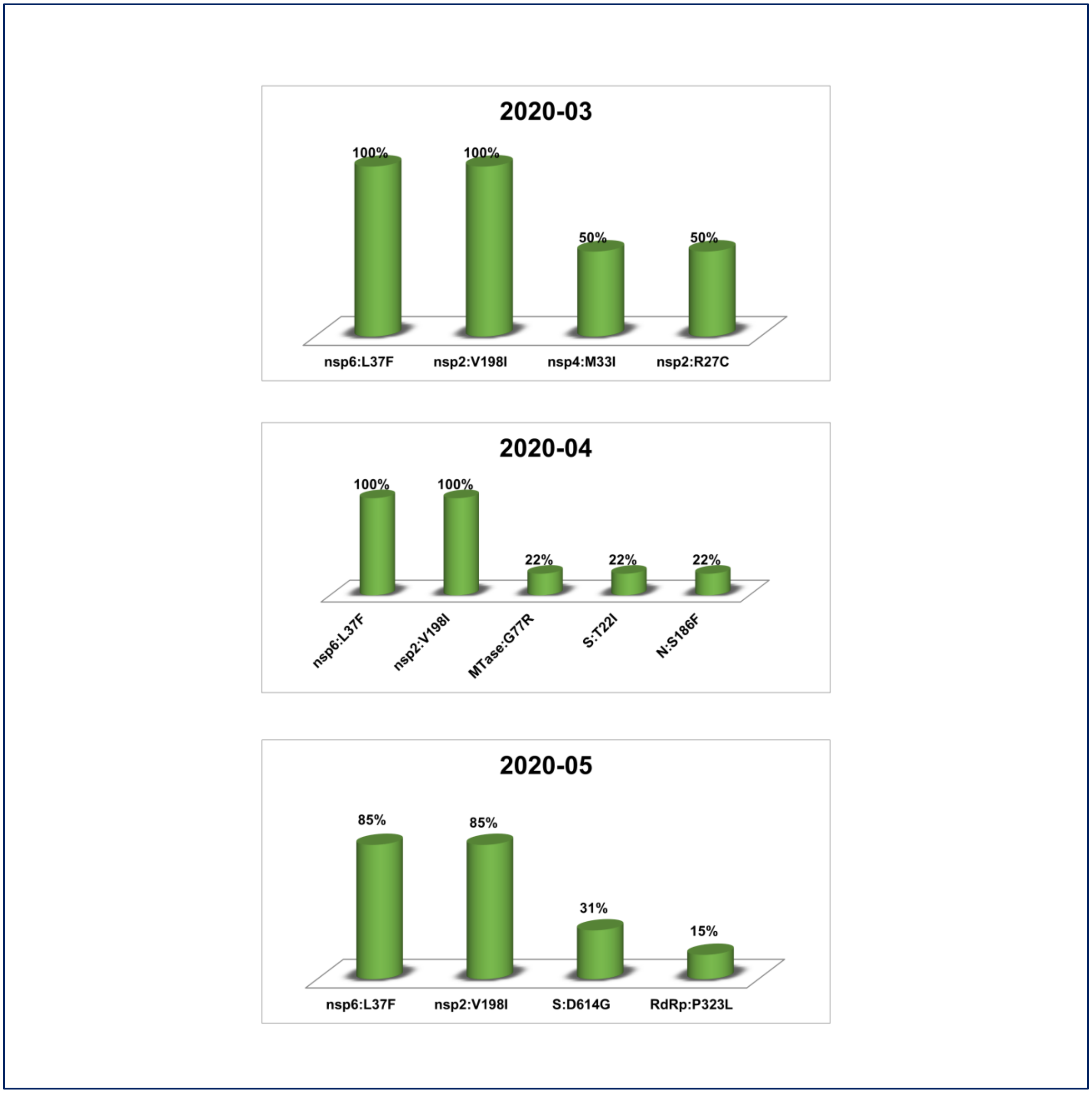

**FigS3.** Top mutations in the second wave of SARS-CoV-2 infection (June-August 2020)

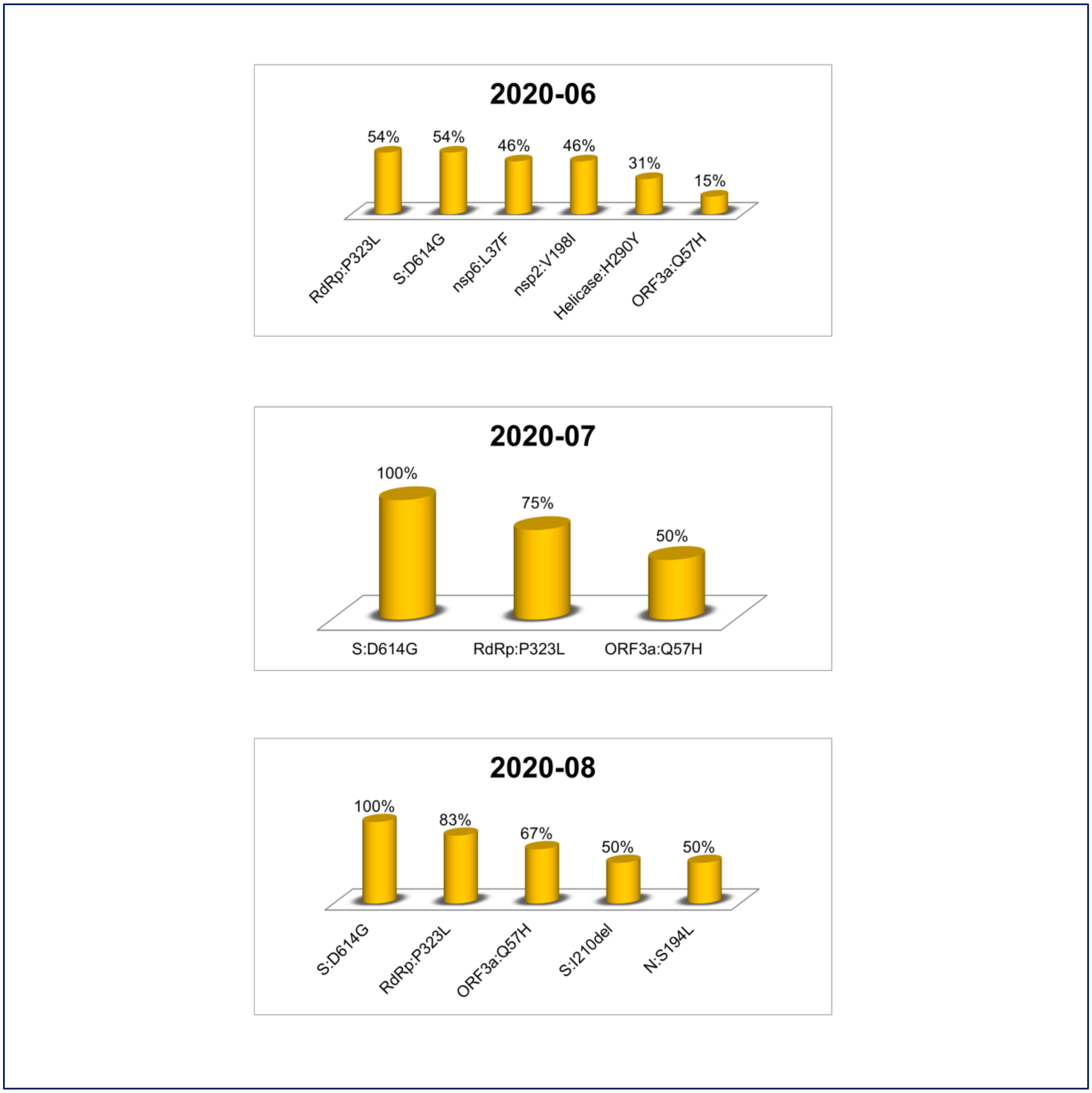

**FigS4.** Top mutations in the third wave of SARS-CoV-2 infection (September-December 2020)

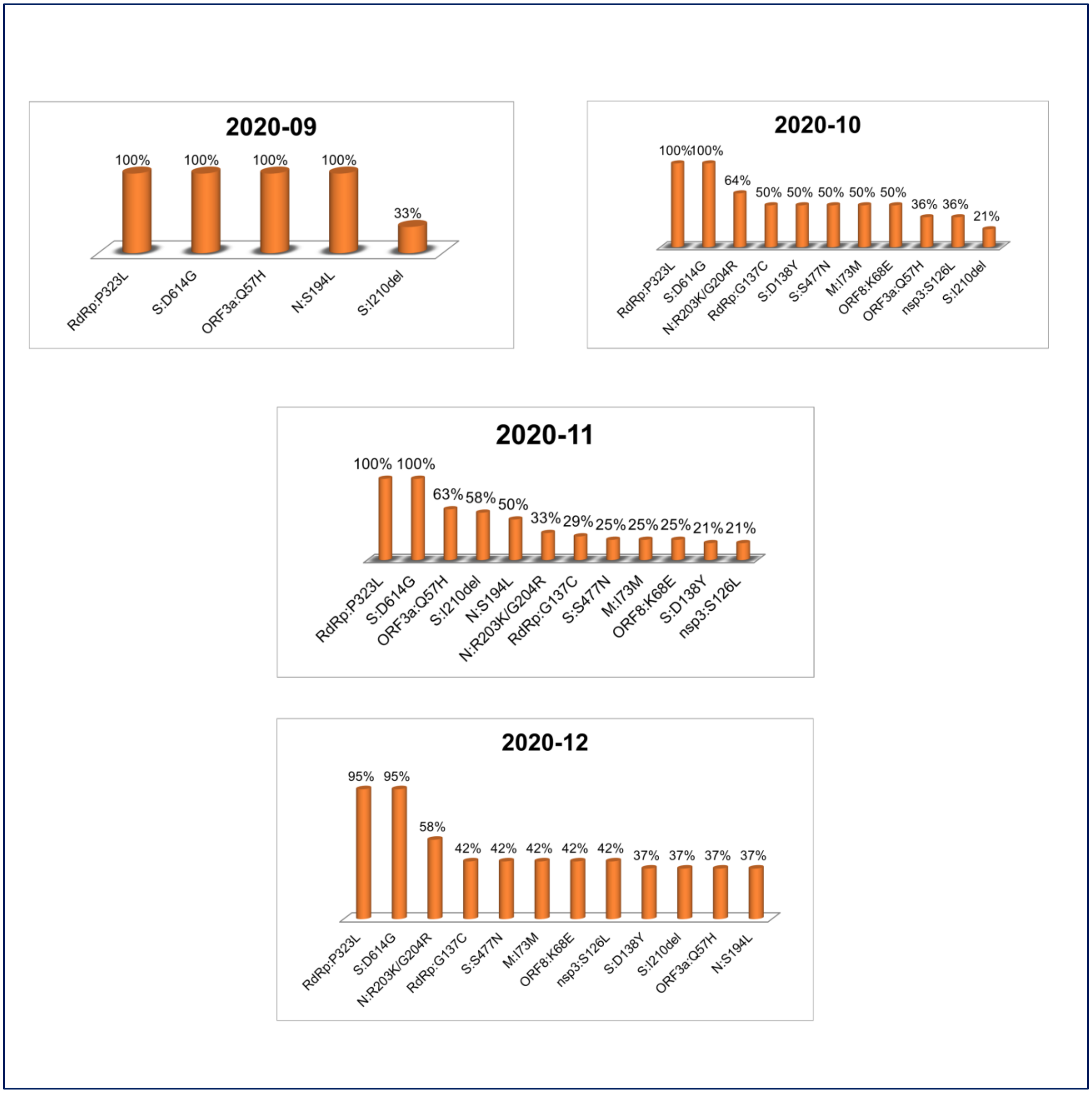

**FigS5.** Top mutations in January-March 2021

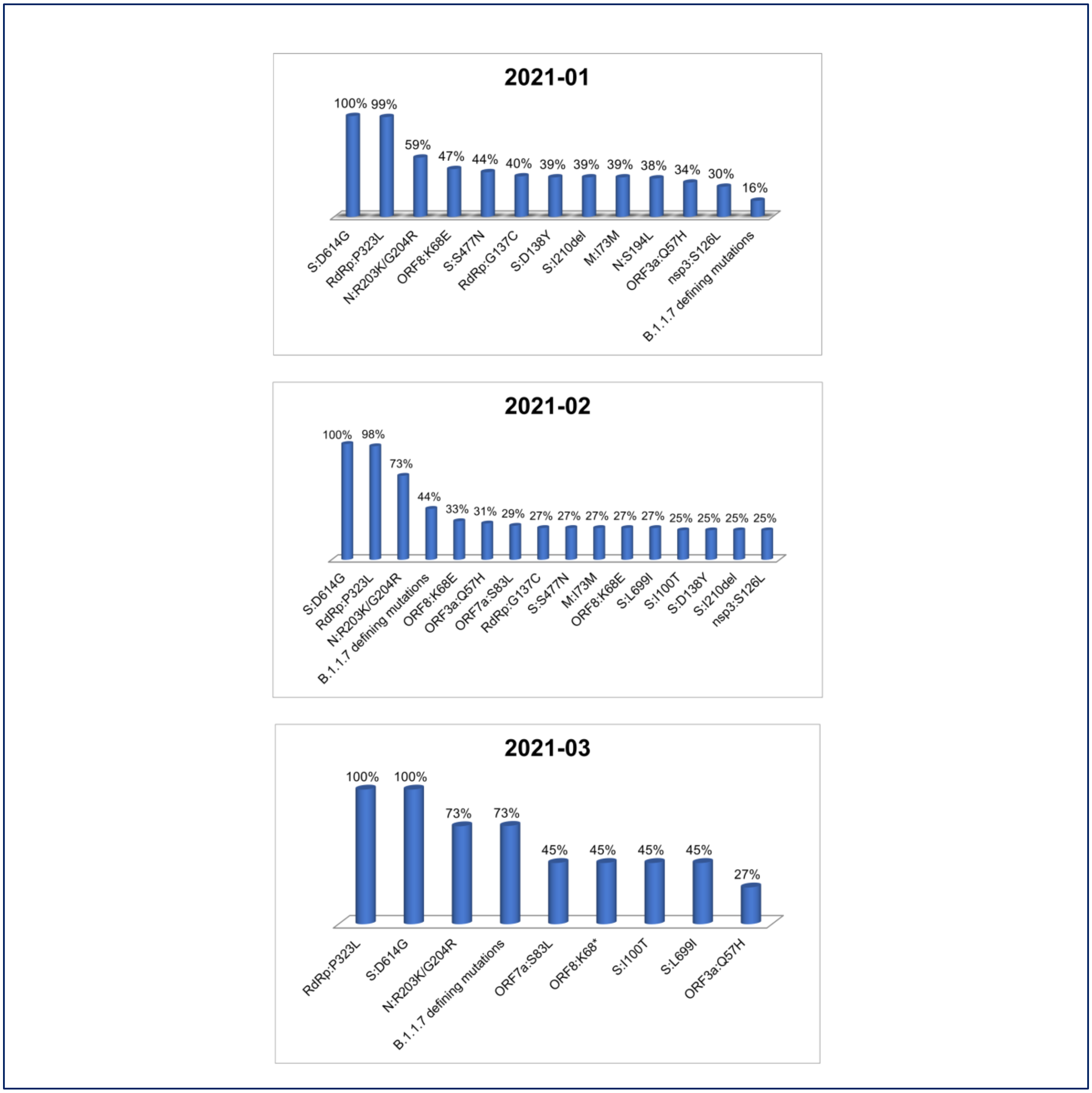

**FigS6.** Top mutations in the fourth wave of SARS-CoV-2 infection (April-May 2021)

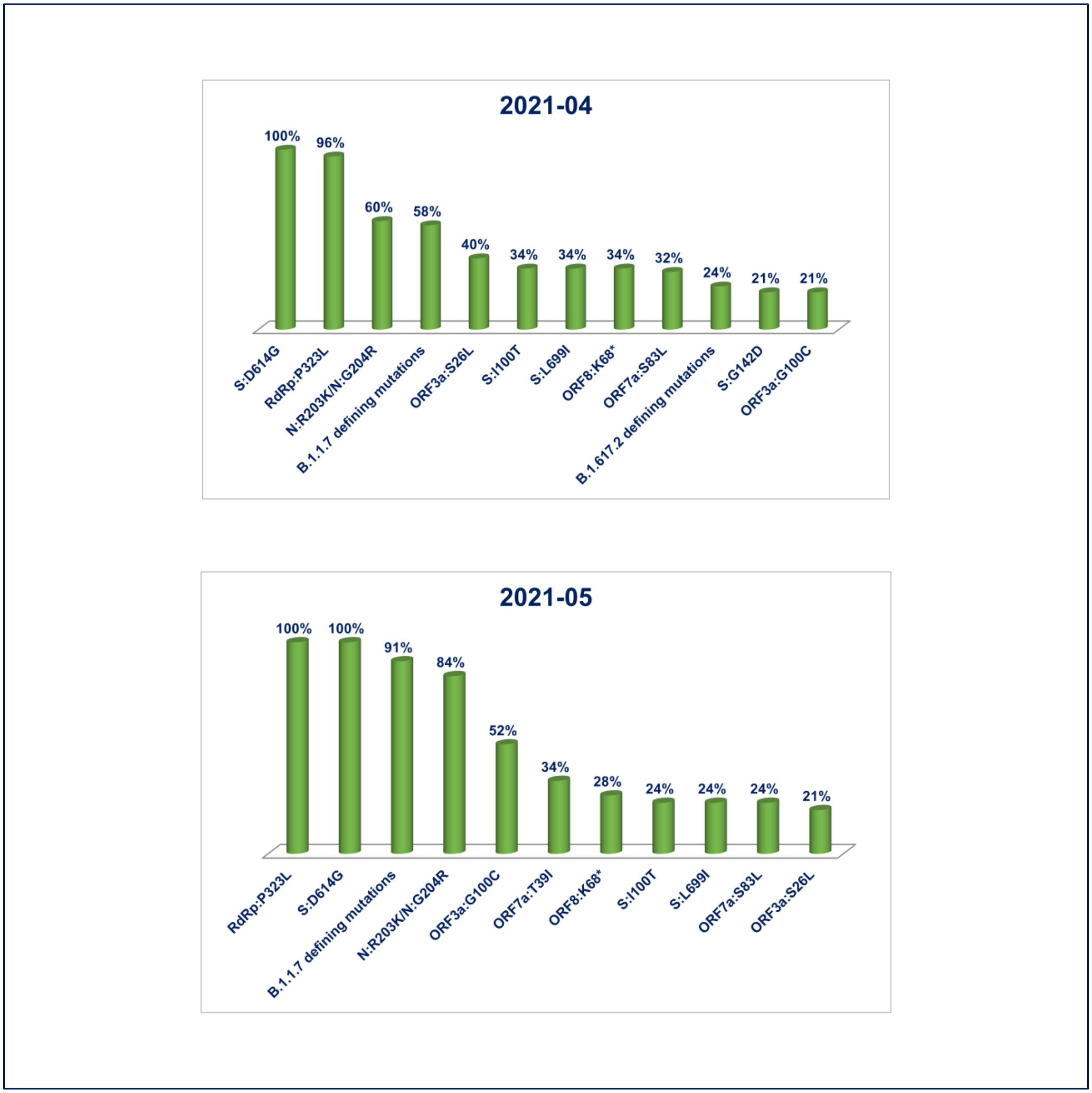
